# Characterisation of populations at risk of sub-optimal dosing of artemisinin-based combination therapy in Africa

**DOI:** 10.1101/2023.05.24.23290481

**Authors:** Abena Takyi, Verena I. Carrara, Prabin Dahal, Marianna Przybylska, Eli Harriss, Genevieve Insaidoo, Karen I. Barnes, Philippe J Guerin, Kasia Stepniewska

**Affiliations:** Centre for Tropical Medicine and Global Health, Nuffield Department of Medicine, University of Oxford, Oxford, United Kingdom; Infectious Diseases Data Observatory (IDDO), Oxford, United Kingdom; WorldWide Antimalarial Resistance Network (WWARN), Oxford, United Kingdom; Department of Child Health, Korle Bu Teaching Hospital, Accra, Ghana; Institute of Global Health, Faculty of Medicine, University of Geneva, Geneva, Switzerland; Royal Infirmary Edinburgh, NHS Lothian, Edinburgh, United Kingdom; The Knowledge Centre, Bodleian Health Care Libraries, University of Oxford, Oxford, OX3 7DQ, UK; Holy family Hospital, Nkawkaw, Eastern Region, Ghana; Division of Clinical Pharmacology, Department of Medicine, University of Cape Town, Cape Town, South Africa

## Abstract

**Introduction:** Selection of resistant malaria strains occurs when parasites are exposed to inadequate antimalarial drug concentrations. The proportion of uncomplicated *falciparum* malaria patients at risk of being sub-optimally dosed with the current World Health Organization (WHO) recommended artemisinin-based combination therapies (ACTs) is unknown. This study aims to estimate this proportion and the excess number of treatment failures (recrudescences) associated with sub-optimal dosing in Sub-Saharan Africa.

**Methods:** Sub-populations at risk of sub-optimal dosing include wasted children <5 years of age; patients with hyperparasitaemia; pregnant women; people living with HIV; and overweight adults. Country-level data on population structure were extracted from openly accessible data sources. Pooled adjusted Hazard Ratios for PCR-confirmed recrudescence were estimated for each risk group from published meta-analyses using fixed-effect meta-analysis.

**Results:** In 2020, of 153.1 million uncomplicated *P. falciparum* malaria patients in Africa, the largest risk groups were the hyperparasitaemic patients (13.2 million, 8.6% of uncomplicated malaria cases) and overweight adults (10.3 million, 6.7% of uncomplicated cases). The excess total number of treatment failures ranged from 323,247 for a 98% baseline ACT efficacy to 1,292,987 for a 92% baseline ACT efficacy.

**Conclusion:** An estimated 1 in nearly 4 people with uncomplicated confirmed *P. falciparum* malaria in Africa are at risk of receiving a sub-optimal antimalarial drug dosing. This increases the risk of antimalarial drug resistance and poses a serious threat to malaria control and elimination efforts. Changes in antimalarial dosing or treatment duration of current antimalarials may be needed and new antimalarials development should ensure sufficient drug concentration levels in these sub-populations that carry a high malaria burden.

## Introduction

In 2021, the World Health Organization (WHO) Africa region alone accounted for approximately 234 of the estimated 247 million malaria cases and 96% of the estimated 619,000 malaria deaths worldwide. Four sub-Saharan Africa countries contributed to about half of the total burden of cases. Increasing investment in malaria control and the scaling up of artemisinin-based combination treatment (ACT) deployment led to a steady decline of 27% in the incidence of malaria cases between 2000 and 2015. Since the number of malaria cases is rising again, most of the increase occurring in the African region [1]. The COVID-19 pandemic seriously disrupted healthcare systems and alongside the direct impact on malaria control programmes, in most endemic countries, access to health care remains challenging for many patients; the 2022 WHO World Malaria Report estimates that an additional 13.4 million cases and 63,000 deaths worldwide were due to disruptions during the pandemic [1].

*Plasmodium falciparum* (*Pf*) is responsible for most cases of severe malaria and the majority of malaria deaths. The continuous reduction in malaria deaths prior to the pandemic, 37% since 2000, persisted despite the increasing number of cases observed. This success might be attributed to the widespread availability of intravenous artesunate followed by an ACT for the treatment of severe malaria [2, 3]. Following the emergence and spread of *Pf* resistant strains to sequential monotherapies, namely chloroquine in the 1960s, followed by sulfadoxine-pyrimethamine in the 1980s [4, 5], and then mefloquine in the 1990s [6], the ACTs became the WHO recommended first-line treatment for uncomplicated *Pf* malaria in 2006 [7]. Since its introduction, artemisinin resistance has been reported in 2007 in Southeast Asia and in Eastern India [8-10]. Resistance to the partner drugs associated with the artemisinin derivatives is of high concern in these regions, leaving very few therapeutic options [11, 12]. With the recent confirmation of independent foci of clinically significant artemisinin resistance emerging on the African continent, specifically in Uganda, Rwanda and Eritrea, and low PCR-adjusted efficacy including in Burkina Faso and Angola, artemisinin and/or partner drug resistance could threaten malaria control and elimination efforts across the continent [13].

Resistance can arise as a consequence of spontaneous changes in the genetic structure of the parasite which provides a competitive advantage allowing it to survive the treatment even when the patient receives recommended doses of ACTs [5]. Another scenario conducive for the selection of resistant parasite strains is inadequate drug exposure [14] or sub-optimal-dosing, a situation where parasites are exposed to an insufficient drug concentration and/or for an inadequate duration to clear the infection [5]. Reduced drug exposure can occur for various reasons including prescription of an inadequate dose (lower than the manufacturer’s recommended dose), poor patient adherence, poor-quality medicines (either sub-standard or falsified medicines with reduced active ingredients), or inadequate absorption (e.g. acute vomiting shortly after drug administration) [15, 16]. These contributory factors may be avoidable. Absorption, distribution or metabolism of the drug, can also differ among specific groups of patients so that taking the same recommended dose in mg/kg body weight can lead to differing drug exposure [14].

As control efforts in Africa result in reduced transmission and case burden of infection, acquired immunity is waning, increasing the risk of more severe forms of the disease as well as resistant strains emerging and surviving in non-immune patients [17]. In the absence of alternatives to artemisinin based antimalarials in the near future, protecting the efficacy of available ACTs by identifying patient groups at high risk of receiving inadequate dosing and finding ways to optimise their treatment is paramount for the success of disease control and elimination.

The current WHO guidelines for malaria [14] identify five groups of population at risk of sub-optimal dosing: (i) malnourished children <5 years of age, (ii) pregnant women, (iii) overweight adults, (iv) patients with uncomplicated hyperparasitaemia, (v) patients co-infected with HIV or TB. WHO states that for these groups “data on antimalarial drug efficacy are still limited and insufficient evidence exists to warrant dose modification”. Close monitoring of these sub-groups is strongly recommended as the risk for treatment failure and/or development of severe malaria with standard drug dosing is increased. However, the current WHO protocol for “methods for surveillance of antimalarial drug efficacy” recommends excluding severely malnourished children, cases of uncomplicated hyperparasitaemia, pregnant women and people living with HIV (PLHIV) from Therapeutic Efficacy Studies (TES) [18, 19]. Consequently, current ACT dosage regimens optimised from trials conducted initially in healthy adults and well-nourished children, must be extrapolated to these excluded populations [20].

This study aims to estimate the proportion of uncomplicated *Pf* malaria cases in endemic African countries at risk of receiving sub-optimal dosing of oral ACTs and to estimate the fraction of treated patients likely to fail treatment because of sub-optimal dosing.

## Methods

African countries with a malaria transmission intensity estimated at one or more cases per 1000 population in 2020 [21] were included. Malaria risk was considered four times higher in rural areas than urban settings based on published entomological inoculation rate estimates. Proportion of risk groups within malaria patient population was assumed to be the same as in the overall country population, but the difference in malaria prevalence between urban and rural areas was accounted for. Levels of malaria endemicity were categorised as hypo-endemic if *Pf* rate in children aged 2-9 years of age was ≤10%; meso-endemic if parasite rate was 11-50%; or hyperendemic if >50%. Malnutrition was defined as wasting (z-score weight-for-height<-2SD), overweight as body-mass index (BMI) ≥ 25kg/m^2^, hyperparasitaemia as >100,000 parasites per microliter. Details of data sources, variables extracted and variables derived are provided in supplementary material p2-8.

As ACT coverage and adherence was not available across all sub-populations, 100% coverage of and adherence with ACTs was assumed.

### Estimation of failure rates on ACTs for sub-population of interest

Absolute and relative estimates of PCR-confirmed recrudescence were extracted from published meta-analyses or systematic literature reviews, searched for on Epistemonikos (supplementary material p9). Two additional systematic reviews were conducted to collate necessary data to support this analysis: one on the efficacy of ACTs in PLHIV (Prospero registration CRD42018089860, study ongoing), and another in non-pregnant, overweight or obese adults (Prospero registration CRD42018090521, available in supplementary material p10-12).

Where available, fixed-effect pooled estimates from meta-analyses’ Hazard Ratios (HR) were calculated by risk group of interest. Otherwise, risk of treatment failure was derived from individual studies and a sensitivity analysis was performed assuming HR range 1.2-2.0. A 2-8% range of hypothetical treatment failure rates in adequately dosed patients was considered, given the current resistance data available from Africa and WHO recommendations to change drug policy if Adequate Clinical and Parasitological Response (ACPR) rate falls below 90% [18].

## Results

### Number of malaria cases

Of 154.6 million confirmed cases, 153.1 million were estimated to be due to uncomplicated malaria of which 37.4 million (24.4%) were in children <5 years of age, 56.1 million (36.6%) in those 5-14 years of age, and 59.6 million (39.0%) in adults >14 years. Country-specific extracted data are provided in supplementary material p12-19. Patients with hyperparasitaemia (13.2 million, 8.6% of uncomplicated malaria cases) and overweight adults (10.3 million, 6.7% of uncomplicated cases) were the largest risk groups in all regions and endemicity areas. Malaria in wasted children was estimated to reach 2.5 million, representing 1.6% of all uncomplicated cases. There were 6.4 million uncomplicated cases in pregnant women, 4.2% of total malaria burden and 10.7% of cases in adult population. The highest proportions of PLHIV and of pregnant women at increased risk of sub-optimal dosing were in East Africa (1.5% and 2.9%, respectively), while wasted children were predominant in meso-endemic regions (2.4% *vs*. 0.1% in hypo-endemic areas), Table 1.

**Table 1.** Number (in millions) of uncomplicated malaria cases per sub-population at increased risk of sub-optimal dosing.

|  |  | Wasted<br>(in <5<br>years) | Pregnancy<br>(in females<br>>14 years) | Overweight<br>(in >14<br>years) | PLHIV<br>(in all ages) | Hyperparasitaemia<br>(in all ages) |
| --- | --- | --- | --- | --- | --- | --- |
| Total | N | 2.5 | 6.4 | 10.3 | 1.9 | 13.2 |
| 41 African<br>countries | % | 1.6 | 4.2 | 6.7 | 1.2 | 8.6 |
| *By region* |  |  |  |  |  |  |
| Northern Africa | N | <0.1 | 0.1 | 0.2 | <0.1 | 0.1 |
| (1 country) | % | 0.0 | 0.1 | 0.1 | 0.0 | 0.1 |
| East Africa | N | 0.5 | 2.9 | 4.7 | 1.5 | 5.2 |
| (15 countries) | % | 0.3 | 1.9 | 3.1 | 1.0 | 3.4 |
| West Africa | N | 1.1 | 2.0 | 3.6 | 0.2 | 4.8 |
| (15 countries) | % | 0.7 | 1.3 | 2.4 | 0.1 | 3.1 |
| Central Africa | N | 0.9 | 1.4 | 1.8 | 0.2 | 3.1 |
| (9 countries) | % | 0.6 | 0.9 | 1.1 | 0.1 | 2.0 |
| Southern Africa | N | <0.1 | <0.1 | <0.1 | <0.1 | <0.1 |
| (1 country) | % | 0.0 | 0.0 | 0.0 | 0.0 | 0.0 |
| *By endemicity<sup>1</sup>* |  |  |  |  |  |  |
| Hypo-endemic | N | 0.1 | 1.0 | 2.0 | 0.3 | 1.5 |
| (16 countries) | % | 0.1 | 0.7 | 1.3 | 0.2 | 0.9 |
| Meso-endemic | N | 2.4 | 5.4 | 8.3 | 1.6 | 11.7 |
| (25 countries) | % | 1.5 | 3.5 | 5.4 | 1.0 | 7.7 |

Percentages are in total of uncomplicated cases. The list of countries by region and by endemicity areas are reported in supplementary material p12-13.
<sup>1</sup> Hypo-endemicity: *Plasmodium falciparum* (*Pf*) prevalence in 2-9 years old <10%; Meso-endemicity: *Pf* prevalence in 2-9 years old 11-50%. No country was reported as hyper-endemic in 2020.
In <5 years: children under 5 years old; in >14 years: adults aged 15 years and older.

Distribution of estimated malaria cases across risk groups varied between countries (Fig. 1 and supplementary material p17-19). The proportion of PLHIV with malaria varied between <0.1 and 4.4% in all countries except Zimbabwe and Namibia, where this sub-population harboured an estimated 7.9 and 8.0% of all uncomplicated *Pf* cases respectively. The proportion of overweight adults varied between 10 and 32% of adults with uncomplicated *Pf* malaria. Proportion of wasted children among children under 5 years with uncomplicated *Pf* malaria was the highest in South Sudan (24%) and Djibouti (30%), (supplementary material p19).

**Fig 1.**
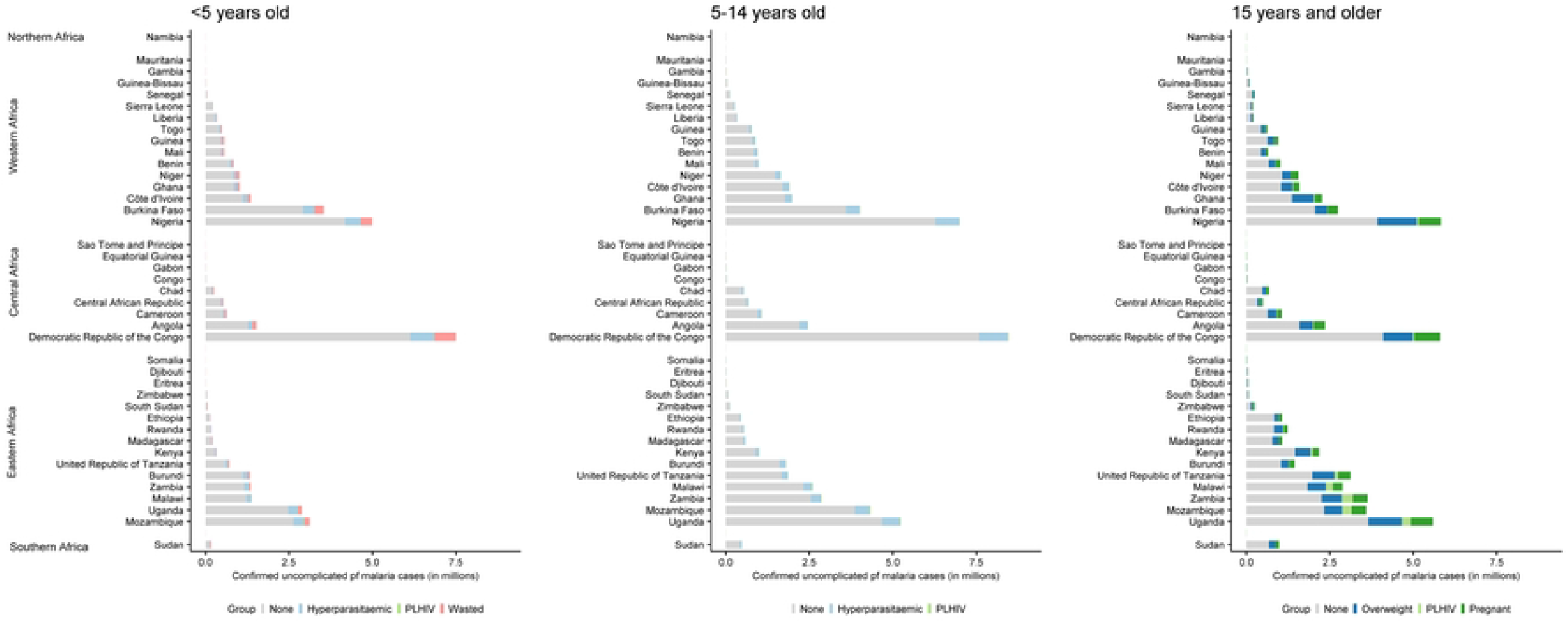
Number (in million) of estimated uncomplicated *Pf* malaria cases by country and region, showing sub-population distribution with increased risk of sub-optimal dosing.

### Number of treatment failures

The systematic review identified five IPD meta-analyses which provided HR estimates for hyperparasitaemic patients (PRISMA flow diagram in supplementary material p9), and individual studies provided estimates for PLHIV (n=4) and malnourished children <5 years of age (n=1), Table 2. No relevant studies were identified for overweight or obese patients (supplementary material p10-12) nor pregnant women.

**Table 2.**
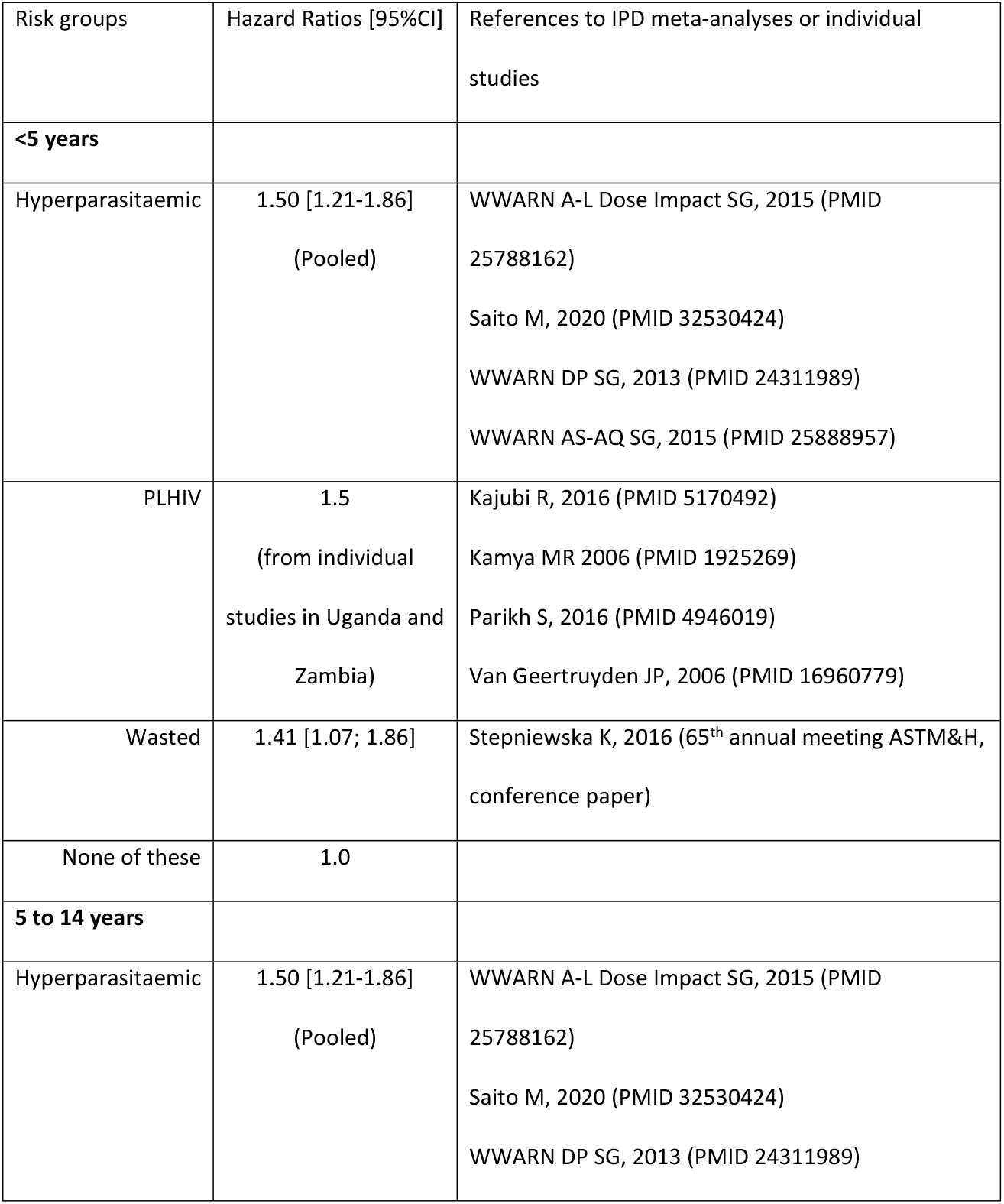
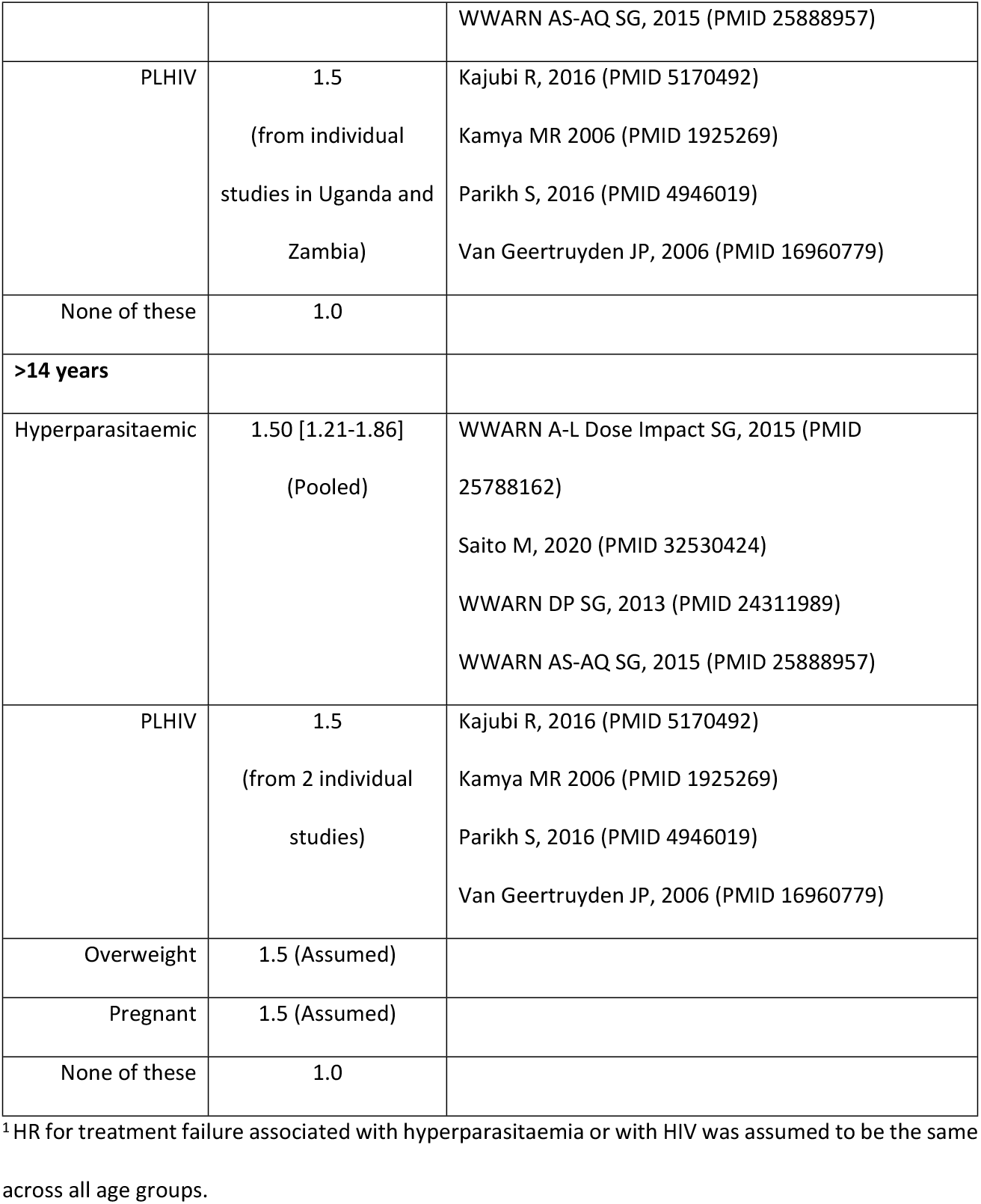
Risk of treatment failure by age group and sub-population at increased risk of sub-optimal dosing used in calculation of the excess number of malaria infections.^1^.

At drug efficacy of 98%, 95% and 92%, the expected number of PCR-corrected treatment failures (recrudescences) were estimated as: 3.1, 7.6 or 12.3 million, and the number of excess rates as 0.4, 1.1 or 1.4 million, respectively (assuming HR=1.5 for pregnant women and overweight patients). The largest contribution to the excess number of treatment failures came from hyperparasitaemic patients (44.2%), (Table 3, Fig. 2, supplementary material p20). Overweight adults, pregnant women, and PLHIV contributed to 23.6%, 14.6%, and 4.7% of excess failures, respectively, which, in a sensitivity analysis, changed to 12.7%, 7.8%, 2.5% (HR=1.2 assumed) and to 33.0%, 20.4%, 6.6% (HR=2.0 assumed), respectively. Wasted children contributed to 12.9% excess failures.

**Table 3:** Excess failures (in millions) estimations in different risk groups, assuming different treatment failure rates and a range of assumed Hazard Ratios (HR) for PLHIV, pregnant women, and overweight adults.

|  | Main Analysis |  |  | Sensitivity analysis |  |  |  |  |  |
| --- | --- | --- | --- | --- | --- | --- | --- | --- | --- |
|  | Assumed HR=1.5<br>for PLHIV, Overweight,<br>Pregnant |  |  | Assumed HR=1.2<br>for for PLHIV,<br>Overweight, Pregnant |  |  | Assumed HR=2<br>for for PLHIV,<br>Overweight, Pregnant |  |  |
| Risk groups | 2%<br>failure | 5%<br>failure | 8%<br>failure | 2%<br>failure | 5%<br>failure | 8%<br>failure | 2%<br>failure | 5%<br>failure | 8%<br>failure |
| **<5 years** |  |  |  |  |  |  |  |  |  |
| Hyperparasitaemic | 0.096 | 0.240 | 0.384 | 0.096 | 0.240 | 0.384 | 0.096 | 0.240 | 0.384 |
| PLHIV | 0.002 | 0.006 | 0.009 | 0.001 | 0.002 | 0.004 | 0.005 | 0.011 | 0.018 |
| Wasted | 0.056 | 0.140 | 0.225 | 0.056 | 0.140 | 0.225 | 0.056 | 0.140 | 0.225 |
| sub-total for <5y | 0.154 | 0.386 | 0.618 | 0.153 | 0.382 | 0.613 | 0.157 | 0.391 | 0.627 |
| **5-14 years** |  |  |  |  |  |  |  |  |  |
| Hyperparasitaemic | 0.057 | 0.143 | 0.228 | 0.057 | 0.143 | 0.228 | 0.057 | 0.143 | 0.228 |
| PLHIV | 0.001 | 0.004 | 0.006 | 0.001 | 0.001 | 0.002 | 0.003 | 0.007 | 0.012 |
| sub-total for 5-14y | 0.058 | 0.147 | 0.234 | 0.058 | 0.144 | 0.230 | 0.060 | 0.150 | 0.240 |
| **>14 years** |  |  |  |  |  |  |  |  |  |
| Hyperparasitaemic | 0.039 | 0.098 | 0.157 | 0.039 | 0.098 | 0.157 | 0.039 | 0.098 | 0.157 |
| PLHIV | 0.017 | 0.042 | 0.067 | 0.007 | 0.017 | 0.027 | 0.034 | 0.084 | 0.135 |
| Overweight | 0.103 | 0.257 | 0.411 | 0.041 | 0.103 | 0.164 | 0.205 | 0.514 | 0.822 |
| Pregnant | 0.064 | 0.159 | 0.255 | 0.025 | 0.064 | 0.102 | 0.127 | 0.318 | 0.509 |
| sub-total for >14y | 0.223 | 0.556 | 0.890 | 0.112 | 0.282 | 0.450 | 0.405 | 1.014 | 1.623 |
| **Overall TOTAL failures** | **0.435** | **1.089** | **1.742** | **0.323** | **0.808** | **1.293** | **0.622** | **1.555** | **2.490** |

**Fig 2.**
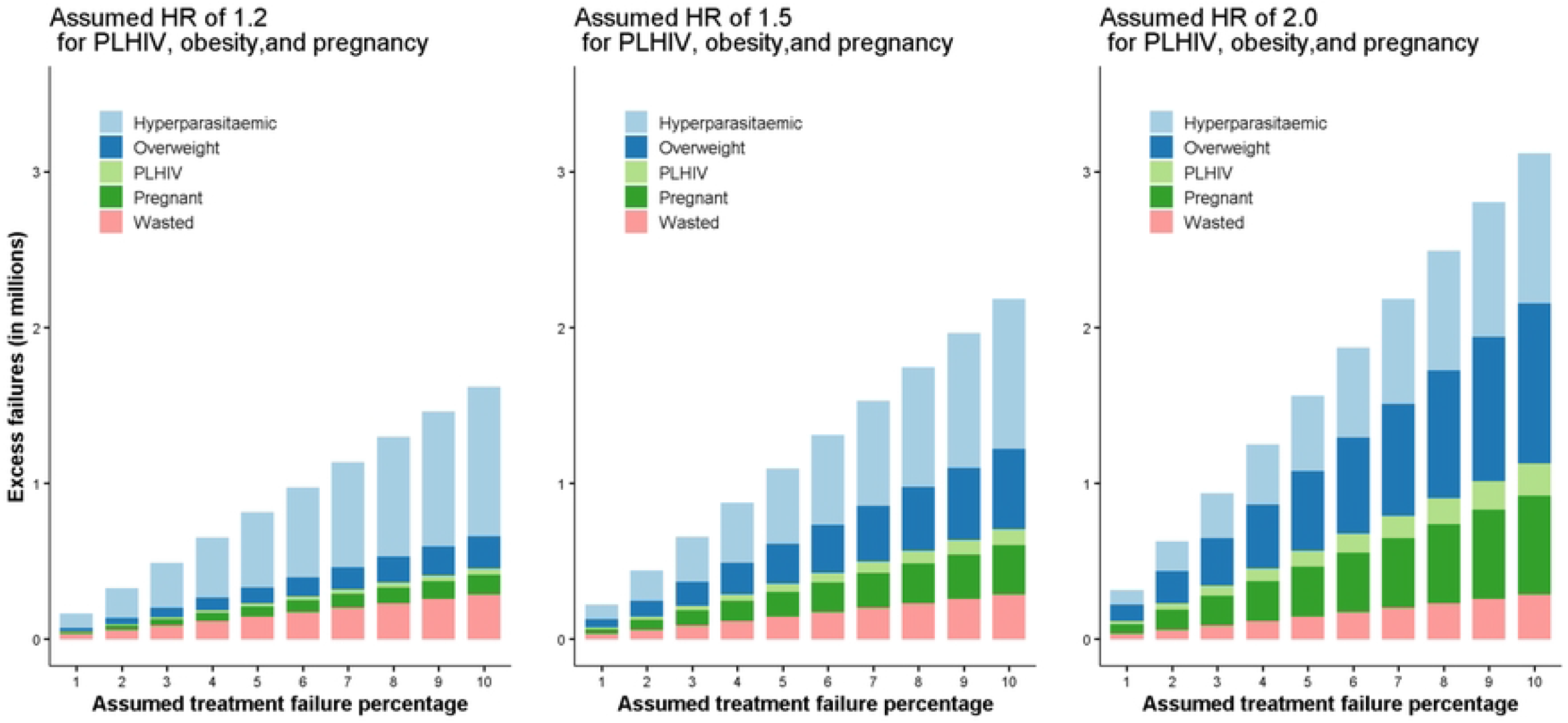
Estimated number of excess treatment failures in millions for different baseline treatment efficacy assuming Hazard Ratio (HR) of 1.2, 1.5 and 2.0 in patients living with HIV (PLHIV), overweight adults, and pregnant women.

Three of the five IPD meta-analyses consistently identified that children <5 years of age with uncomplicated malaria but without the above-mentioned risk factors were also at the increased risk of treatment failure when compared to adults (HR 2.68 [95%CI: 1.87-3.85], supplementary material p21-24) and were associated with an estimated additional number of failure rates of 1.050 - 4.202 million for ACPRs between 98% and 92%.

## Discussion

This study estimated that nearly one in four uncomplicated *Pf* malaria patients in Africa are within a sub-population of patients considered at risk of sub-optimal ACT dosing by the WHO [1]. We estimated that excess annual treatment failures could range between 0.32 to 1.29 million, 0.44 to 1.09 million and 0.62 to 2.49 million individuals in the five identified sub-populations with an ACPR of 98%, 95% and 92%, respectively.

Until optimised dosage regimens are defined for these groups, the close monitoring of treatment response in all those at risk of sub-optimal dosing will become paramount to successfully limit the emergence and spread of artemisinin- and partner drug-resistant parasite strains on the African continent. This is especially important at a time when clinically significant artemisinin resistance has been confirmed in at least three African countries [13] and when acquired immunity is waning in regions successfully controlling the overall malaria burden [17]; and is part of the new WHO strategy to minimize the threat and impact of antimalarial drug resistance in Africa [13].

Patients with uncomplicated hyperparasitaemia accounted for 13.2 million, or 8.6%, of estimated uncomplicated malaria cases and are the largest contributor of estimated excess treatment failures. In this study uncomplicated hyperparasitaemia was defined as >100,000 parasites/μL, based on two meta-analyses defining this as the threshold for an increased risk of treatment failure [22, 23] and its proportion based on a meta-analysis on 56,000 individual patients’ data that included 29 African countries in low, moderate, and high malaria transmission areas [24]. This proportion however may be an underestimation as patients with this level of parasitaemia are often excluded from uncomplicated malaria clinical trials [19]. Patients with uncomplicated hyperparasitaemia and without other clinical signs of severity are an important reservoir of de-novo resistance [25]. Additionally, inadequate treatment may aggravate the patient’s clinical condition and increase risk of death [26]. Although severely ill hyperparasitaemic patients are likely to be hospitalised and treated parentally, recognising a patient with isolated uncomplicated hyperparasitaemia is challenging as diagnosis is usually made by qualitative malaria rapid diagnostic tests and without microscopic confirmation of parasite density. These patients are thus likely to receive a standard oral ACT dosage regimen that may be insufficient to reduce their high parasite biomass thus increasing the risk of recrudescence [25]. Once the diagnosis of uncomplicated hyperparasitaemia is made then the treatment remains problematic as evidence to date to support, e.g. increasing the ACT duration, is insufficient [14, 27]. Malaria and undernutrition often coincide in Africa, where approximately 1 in 3 children under 5 years of age are underweight, supplementary material p25. The risk of malaria and treatment failure according to nutritional status remains complex [28]. Furthermore, malnutrition may worsen the severity of malaria and increase the risk of malaria deaths [29] and acutely undernourished (wasted) children are at an increased risk of ACT treatment failure [30]. This is reflected within the current WHO malaria treatment guidelines when referring to children “malnourished” as being at risk of sub-optimal dosing. In 2019, a year prior to the COVID-19 pandemic, an estimated 12.7 million children <5 years of age in Africa were acutely malnourished, of whom 3.5 million were considered severely wasted (weight-for-height Z-score <-3SD) and at higher risk of infection, complications and death [31]. The many social, economic, and health-related disruptions triggered by the COVID-19 pandemic alongside the current food shortage due to the war in Ukraine aggravate the nutritional status of an additional 1.46 million children in Africa [32]. The present study estimates that 2.5 million moderately (weight-for-height Z-score <-2SD) wasted children <5 years of age suffer from uncomplicated malaria; unless they present with danger signs or complications from their malnutrition status, these children are likely to be treated with ACTs including those in nutrition rehabilitation [33]. There is strong evidence from studies conducted in Mali and Niger that severely wasted children, treated with a full course of artemether-lumefantrine and high-fat nutritional supplements, have decreased drug exposure and a higher risk of reinfection compared to those who are well-nourished [34]. Importantly, even mild wasting (weight-for-height Z-score <-1SD) increases the risk of treatment failure to the above estimates underestimate the total effects of wasting on ACT treatment failure [30]. Furthermore, in the WWARN individual pharmacokinetic-pharmacodynamic data analysis of patients treated with artemether-lumefantrine, underweight (weight-for-age Z score <-2 SD) children under 3 years of age had a 23% [95%CI 1; 41] lower day 7 lumefantrine concentration [35] and underweight African children <3 years of age had a higher risk of treatment failure (HR 1.66 [95%CI 1.05; 2.63]) compared to adequately-nourished children of the same age [23]. Improving our understanding of the complex interactions between nutritional status, antimalarial drug absorption and ACT efficacy is paramount to improve clinical management of these patients and avoid preventable treatment failures and increasing antimalarial resistance.

In this study, persons who are overweight accounted for an estimated 10.3 million, or 6.7% of all estimated uncomplicated malaria cases, the second largest risk group. The pharmacological profile of lipophilic antimalarial drugs in overweight or obese people may be altered. One small recent pharmacokinetic study on healthy males showed non-significantly lowered artemether-lumefantrine plasma drug concentrations with higher body-weight, but was likely underpowered with only 7 overweight and 3 obese participants included [36]. Publications evaluating their risks of sub-optimal dosing [37], recrudescence or even severity are still too sparse to provide reliable estimates of effect, which is expected to vary with both the degree of obesity, antimalarial used, and level of immunity among adults enrolled. One study conducted in Sweden retrospectively reviewed medical charts of patients hospitalised with falciparum malaria and concluded that median body mass index (BMI) in patients with severe malaria was significantly higher (29.3 kg/m^2^) than for those with uncomplicated malaria, concluding obesity (BMI ≥30 kg/m^2^) was significantly associated with severe malaria at diagnosis [38]. A study by Hatz *et al*. in 165 non-immune adults reported a decreased artemether-lumefantrine day-28 parasitological cure rate (93.4% [95%CI 85.3; 97.8] in patients ≥65 kg compared to those <65 kg (100% [95%CI 92.5; 100]) [39]. In principle, dosing of ACTs should be based on a target mg/kg body weight dose, but ACTs are mostly available as pre-packaged treatments based on a single adult weight-band (e.g. artemether-lumefantrine dosage is identical for anyone weighing ≥35 kg) [14]. Increasing the treatment dose or prolonging the treatment regimen [40] for overweight patients could be feasible; however, it may be challenging in some primary health care contexts. As malaria transmission intensity decreases, the age distribution of malaria morbidity and mortality burden expands, with increased prevalence of malaria in the adult population. In parallel an increase in the prevalence of overweight/obesity in African adults has also been observed [41-43]. Therefore, improving diagnosis and treatment in older age groups remains relevant to advance elimination and delay resistance [44]; thus overweight adults should be actively included in dose optimization studies to provide data on this important population.

PLHIV could contribute to 1.2% of all estimated uncomplicated malaria cases and between 2.6% and 6.6% of estimated excess failures. As antiretroviral therapy (ART) coverage increases [45] together with a shift towards dolutegravir-based ARTs that have fewer drug-drug interactions [46], PLHIV may become less at risk of sub-optimal ACT dosing with standard 3-day regimen; this risk remains however for those receiving rifampicin-based tuberculosis treatment or efavirenz-based ARTs [47, 48]. Furthermore, PLHIV have higher parasites densities and children infected with HIV have been reported having slower parasite clearance than HIV-free children [49]. A recent review on the role of HIV infection on malaria transmission suggests a higher risk of re-infection in population infected with HIV-1 [50].

The current WHO Malaria guidelines for treating uncomplicated malaria in pregnancy recommend that artemether-lumefantrine should be used in all trimesters [14]. However, artemether-lumefantrine, the most widely used ACT in Africa, had a lower PCR-corrected cure rate compared to other ACTs in a large IPD meta-analysis evaluating the efficacy and tolerability of ACTs in pregnancy [51], which could be attributed to changes in the pharmacokinetics of lumefantrine during pregnancy resulting in lower drug concentration compared to non-pregnant population [52]. Longer artemether-lumefantrine regimens have been tested in Thailand and in the Democratic Republic of Congo, with a higher Day-7 lumefantrine concentration compared to the standard 3-day regimen but did not show increase ACPRs [53]. Further studies to optimise antimalarial drug treatment in pregnancy are needed, as are harmonised antimalarial therapeutic efficacy assessments in pregnancy studies [54].

## Study limitations and assumptions

This study provides an estimate of the significant magnitude of the population at risk of sub-optimal dosing living in 41 African countries; those estimates are based on the latest malaria and population data openly available and are derived from several sources with some assumptions. Assumptions made to evaluate the number of treatment failure and the proportion of excess treatment failure for each sub-population evaluated. Estimates have been calculated assuming an equal risk for everyone in a population sub-group and an equal risk by age category within that sub-group. The estimated incidence of severe malaria cases from 2015 was applied to calculate uncomplicated episodes from the 2020 total malaria data reported by WHO although malaria trends were decreasing until 2019.

Recent IPD meta-analysis were not available for each population category nor were exclusively evaluating treatment failure risk in Africa. Risks of treatment failure associated with multiple factors could not be evaluated (e.g., hyperparasitaemia in pregnancy). Because sub-group populations and malaria endemicity levels were extracted at country level, granularity of risk may have been lost including level of transmission or impact of seasonality. We did not account for the quality of antimalarials (either substandards or falsified), the impact of other co-morbidities on the drug absorption, the impact of drugs other than antiretrovirals e.g. antituberculosis drugs or the true adherence to the treatment. We believe that the majority of our assumptions are likely to underestimate the true overall impact of under-dosing, so provide a “best case” scenario.

## Conclusion

This study estimates that nearly 1 in 4 people with uncomplicated confirmed malaria in Africa are at risk of sub-optimal antimalarial drug dosing. This is the first attempt to quantify this issue, which poses a serious threat to malaria control efforts. Adequate antimalarial drug dosing is essential for both maximising cure rates and the prevention or delay of resistance emergence or its expansion. Optimised drug dosing or longer treatment duration of currently used ACTs may be needed in those at risk of sub-optimal antimalarial drug dosing. The largest contribution to the excess number of treatment failures came from hyperparasitaemic patients. A malaria diagnosis that includes a quantitative or semi-quantitative parasite count at all levels of health care would be of great public health value to identify patients with uncomplicated hyperparasitaemia who should receive an adapted treatment. New antimalarials should be evaluated to provide sufficient drug concentrations not only in otherwise healthy adults, but also to all at risk sub-populations that carry a high malaria burden.

## Data Availability

Data from cited openly available sources were accessed and data used for the analysis are available in the supplementary materials.

## Acknowledgments

The authors would like to thank Jamie T. Griffin and colleagues for sharing the data used in their model for estimating the proportion of malaria cases in each age group according to the different types of malaria transmission (Figure 3c from the manuscript entitled Griffin, J.T., N.M. Ferguson, and A.C. Ghani, *Estimates of the changing age-burden of Plasmodium falciparum malaria disease in sub-Saharan Africa*. Nat Commun, 2014. **5**: p. 3136). This study is supported by the Bill & Melinda Gates Foundation (grant INV-004713).

## Supplementary material

Supplementary material is available as one appendix

## Notes

### Competing Interest Statement

The authors have declared no competing interest.

### Funding Statement

Bill & Melinda Gates Foundation (grant INV-004713) and the University of Oxford. The funders of the study had no role in the study design, evidence synthesis, writing of the manuscript or the decision to submit it for publication.

### Author Declarations

Patient consent for publication: Not required. Ethics approval: Not required.

## References

1. World malaria report 2022. Geneva: World Health Organization, 2022.

2. Thwing J, Eisele TP, Steketee RW. Protective efficacy of malaria case management and intermittent preventive treatment for preventing malaria mortality in children: a systematic review for the Lives Saved Tool. BMC Public Health. 2011;11 Suppl 3:S14. Epub 2011/04/29. doi: 10.1186/1471-2458-11-S3-S14. PubMed PMID: 21501431; PubMed Central PMCID: PMCPMC3231887.

3. White NJ, Pukrittayakamee S, Hien TT, Faiz MA, Mokuolu OA, Dondorp AM. Malaria. Lancet. 2014;383(9918):723–35. doi: 10.1016/S0140-6736(13)60024-0. PubMed PMID: 23953767.

4. Roper C, Pearce R, Nair S, Sharp B, Nosten F, Anderson T. Intercontinental spread of pyrimethamine-resistant malaria. Science. 2004;305(5687):1124. PubMed PMID: 15326348.

5. White NJ. Antimalarial drug resistance. J Clin Invest. 2004;113(8):1084–92. Epub 2004/04/16. doi: 10.1172/JCI21682. PubMed PMID: 15085184; PubMed Central PMCID: PMCPMC385418.

6. Mockenhaupt FP. Mefloquine resistance in Plasmodium falciparum. Parasitol Today. 1995;11(7):248–53. Epub 1995/07/01. doi: 10.1016/0169-4758(95)80201-0. PubMed PMID: 15275335.

7. WHO briefing on Malaria Treatment Guidelines and artemisinin monotherapies. Geneva: World Health Organization, 2006.

8. Dondorp AM, Nosten F, Yi P, Das D, Phyo AP, Tarning J, et al. Artemisinin resistance in Plasmodium falciparum malaria. N Engl J Med. 2009;361(5):455–67. doi: 10.1056/NEJMoa0808859. PubMed PMID: 19641202; PubMed Central PMCID: PMCPMC3495232.

9. Mishra N, Kaitholia K, Srivastava B, Shah NK, Narayan JP, Dev V, et al. Declining efficacy of artesunate plus sulphadoxine-pyrimethamine in northeastern India. Malar J. 2014;13:284. Epub 20140722. doi: 10.1186/1475-2875-13-284. PubMed PMID: 25052385; PubMed Central PMCID: PMCPMC4127069.

10. Tun KM, Imwong M, Lwin KM, Win AA, Hlaing TM, Hlaing T, et al. Spread of artemisinin-resistant Plasmodium falciparum in Myanmar: a cross-sectional survey of the K13 molecular marker. Lancet Infect Dis. 2015;15(4):415–21. Epub 20150220. doi: 10.1016/S1473-3099(15)70032-0. PubMed PMID: 25704894; PubMed Central PMCID: PMCPMC4374103.

11. Amaratunga C, Lim P, Suon S, Sreng S, Mao S, Sopha C, et al. Dihydroartemisinin-piperaquine resistance in Plasmodium falciparum malaria in Cambodia: a multisite prospective cohort study. Lancet Infect Dis. 2016;16(3):357–65. Epub 20160108. doi: 10.1016/S1473-3099(15)00487-9. PubMed PMID: 26774243; PubMed Central PMCID: PMCPMC4792715.

12. Shrestha B, Shah Z, Morgan AP, Saingam P, Chaisatit C, Chaorattanakawee S, et al. Distribution and Temporal Dynamics of Plasmodium falciparum Chloroquine Resistance Transporter Mutations Associated With Piperaquine Resistance in Northern Cambodia. J Infect Dis. 2021;224(6):1077–85. doi: 10.1093/infdis/jiab055. PubMed PMID: 33528566; PubMed Central PMCID: PMCPMC8448439.

13. Strategy to respond to antimalarial drug resistance in Africa. Geneva: World Health Organization, 2022.

14. WHO Guidelines for malaria, 3 June 2022. Geneva: World Health Organization, 2022.

15. Barnes KI, Watkins WM, White NJ. Antimalarial dosing regimens and drug resistance. Trends Parasitol. 2008;24(3):127–34. Epub 20080211. doi: 10.1016/j.pt.2007.11.008. PubMed PMID: 18262470.

16. Leslie T, Kaur H, Mohammed N, Kolaczinski K, Ord RL, Rowland M. Epidemic of Plasmodium falciparum malaria involving substandard antimalarial drugs, Pakistan, 2003. Emerg Infect Dis. 2009;15(11):1753–9. doi: 10.3201/eid1511.090886. PubMed PMID: 19891862; PubMed Central PMCID: PMCPMC2857251.

17. Pongtavornpinyo W, Yeung S, Hastings IM, Dondorp AM, Day NP, White NJ. Spread of antimalarial drug resistance: mathematical model with implications for ACT drug policies. Malar J. 2008;7:229. Epub 2008/11/04. doi: 10.1186/1475-2875-7-229. PubMed PMID: 18976503; PubMed Central PMCID: PMCPMC2585590.

18. Methods for surveillance of antimalarial drug efficacy. Geneva: World Health Organization, 2009.

19. Arena L, Zanamwe M, Halleux CM, Carrara V, Angus BJ, Ariana P, et al. Malaria patient spectrum representation in therapeutic clinical trials of uncomplicated malaria: a scoping review of the literature. Malar J. 2023;22(1):50. Epub 20230210. doi: 10.1186/s12936-023-04441-5. PubMed PMID: 36765317; PubMed Central PMCID: PMCPMC9913008.

20. Tarning J, Zongo I, Some FA, Rouamba N, Parikh S, Rosenthal PJ, et al. Population pharmacokinetics and pharmacodynamics of piperaquine in children with uncomplicated falciparum malaria. Clin Pharmacol Ther. 2012;91(3):497–505. Epub 2012/01/20. doi: 10.1038/clpt.2011.254. PubMed PMID: 22258469; PubMed Central PMCID: PMCPMC3736305.

21. Mendis K, Rietveld A, Warsame M, Bosman A, Greenwood B, Wernsdorfer WH. From malaria control to eradication: The WHO perspective. Trop Med Int Health. 2009;14(7):802–9. Epub 2009/06/06. doi: 10.1111/j.1365-3156.2009.02287.x. PubMed PMID: 19497083.

22. WorldWide Antimalarial Resistance Network AS-AQ Study Group. The effect of dosing strategies on the therapeutic efficacy of artesunate-amodiaquine for uncomplicated malaria: a meta-analysis of individual patient data. BMC Med. 2015;13:66. Epub 20150331. doi: 10.1186/s12916-015-0301-z. PubMed PMID: 25888957; PubMed Central PMCID: PMCPMC4411752.

23. Worldwide Antimalarial Resistance Network A-L Dose Impact Study Group. The effect of dose on the antimalarial efficacy of artemether-lumefantrine: a systematic review and pooled analysis of individual patient data. Lancet Infect Dis. 2015;15(6):692–702. Epub 20150316. doi: 10.1016/S1473-3099(15)70024-1. PubMed PMID: 25788162; PubMed Central PMCID: PMCPMC4632191.

24. WorldWide Antimalarial Resistance Network Falciparum Haematology Study Group. Haematological consequences of acute uncomplicated falciparum malaria: a WorldWide Antimalarial Resistance Network pooled analysis of individual patient data. BMC Med. 2022;20(1):85. Epub 20220307. doi: 10.1186/s12916-022-02265-9. PubMed PMID: 35249546.

25. White NJ, Pongtavornpinyo W, Maude RJ, Saralamba S, Aguas R, Stepniewska K, et al. Hyperparasitaemia and low dosing are an important source of anti-malarial drug resistance. Malar J. 2009;8:253. Epub 2009/11/13. doi: 10.1186/1475-2875-8-253. PubMed PMID: 19906307; PubMed Central PMCID: PMCPMC2784792.

26. Dondorp AM, Lee SJ, Faiz MA, Mishra S, Price R, Tjitra E, et al. The relationship between age and the manifestations of and mortality associated with severe malaria. Clin Infect Dis. 2008;47(2):151–7. Epub 2008/06/07. doi: 10.1086/589287. PubMed PMID: 18533842.

27. Sowunmi A, Adedeji AA, Fateye BA, Babalola CP. Plasmodium falciparum hyperparasitaemia in children. Risk factors, treatment outcomes, and gametocytaemia following treatment. Parasite. 2004;11(3):317–23. Epub 2004/10/20. doi: 10.1051/parasite/2004113317. PubMed PMID: 15490757.

28. Das D, Grais RF, Okiro EA, Stepniewska K, Mansoor R, van der Kam S, et al. Complex interactions between malaria and malnutrition: a systematic literature review. BMC Med. 2018;16(1):186. Epub 2018/10/30. doi: 10.1186/s12916-018-1177-5. PubMed PMID: 30371344; PubMed Central PMCID: PMCPMC6205776.

29. Ferreira E, Alexandre MA, Salinas JL, de Siqueira AM, Benzecry SG, de Lacerda MV, et al. Association between anthropometry-based nutritional status and malaria: a systematic review of observational studies. Malar J. 2015;14:346. Epub 2015/09/18. doi: 10.1186/s12936-015-0870-5. PubMed PMID: 26377094; PubMed Central PMCID: PMCPMC4574180.

30. Stepniewska K. Assessing the impact of malnutrition on the treatment outcome of artemisinin-based combination therapy in uncomplicated plasmodium falciparum malaria. ASTMH 65th Annual Meeting; Atlanta, Georgia USA 2016. p. 467.

31. UNICEF, WHO, World Bank Group: Joint malnutrition estimates 2020 [cited 2022 August, 6]. Available from: https://www.who.int/publications/i/item/jme-2020-edition.

32. Headey D, Heidkamp R, Osendarp S, Ruel M, Scott N, Black R, et al. Impacts of COVID-19 on childhood malnutrition and nutrition-related mortality. Lancet. 2020;396(10250):519–21. Epub 2020/07/31. doi: 10.1016/S0140-6736(20)31647-0. PubMed PMID: 32730743; PubMed Central PMCID: PMCPMC7384798.

33. Akparibo R, Harris J, Blank L, Campbell MJ, Holdsworth M. Severe acute malnutrition in children aged under 5 years can be successfully managed in a non-emergency routine community healthcare setting in Ghana. Matern Child Nutr. 2017;13(4). Epub 2017/02/12. doi: 10.1111/mcn.12417. PubMed PMID: 28185414; PubMed Central PMCID: PMCPMC6865978.

34. Chotsiri P, Denoeud-Ndam L, Baudin E, Guindo O, Diawara H, Attaher O, et al. Severe Acute Malnutrition Results in Lower Lumefantrine Exposure in Children Treated With Artemether-Lumefantrine for Uncomplicated Malaria. Clin Pharmacol Ther. 2019;106(6):1299–309. Epub 2019/06/04. doi: 10.1002/cpt.1531. PubMed PMID: 31152555; PubMed Central PMCID: PMCPMC6896236.

35. WorldWide Antimalarial Resistance Network Lumefantrine PK-PD Study Group. Artemetherlumefantrine treatment of uncomplicated Plasmodium falciparum malaria: a systematic review and meta-analysis of day 7 lumefantrine concentrations and therapeutic response using individual patient data. BMC Med. 2015;13:227. Epub 20150918. doi: 10.1186/s12916-015-0456-7. PubMed PMID: 26381375; PubMed Central PMCID: PMCPMC4574542.

36. Sugiarto SR, Page-Sharp M, Drinkwater JJ, Davis WA, Salman S, Davis TME. The pharmacokinetic properties of the antimalarial combination therapy artemether-lumefantrine in normal weight, overweight and obese healthy male adults. Int J Antimicrob Agents. 2021:106482. Epub 20211121. doi: 10.1016/j.ijantimicag.2021.106482. PubMed PMID: 34818520.

37. Roseau JB, Pradines B, Paleiron N, Vedy S, Madamet M, Simon F, et al. Failure of dihydroartemisinin plus piperaquine treatment of falciparum malaria by under-dosing in an overweight patient. Malar J. 2016;15:479. Epub 2016/09/21. doi: 10.1186/s12936-016-1535-8. PubMed PMID: 27646822; PubMed Central PMCID: PMCPMC5028982.

38. Wyss K, Wangdahl A, Vesterlund M, Hammar U, Dashti S, Naucler P, et al. Obesity and Diabetes as Risk Factors for Severe Plasmodium falciparum Malaria: Results From a Swedish Nationwide Study. Clin Infect Dis. 2017;65(6):949–58. Epub 2017/05/17. doi: 10.1093/cid/cix437. PubMed PMID: 28510633; PubMed Central PMCID: PMCPMC5848256.

39. Hatz C, Soto J, Nothdurft HD, Zoller T, Weitzel T, Loutan L, et al. Treatment of acute uncomplicated falciparum malaria with artemether-lumefantrine in nonimmune populations: a safety, efficacy, and pharmacokinetic study. Am J Trop Med Hyg. 2008;78(2):241–7. Epub 2008/02/08. PubMed PMID: 18256423.

40. Tun KM, Jeeyapant A, Myint AH, Kyaw ZT, Dhorda M, Mukaka M, et al. Effectiveness and safety of 3 and 5 day courses of artemether-lumefantrine for the treatment of uncomplicated falciparum malaria in an area of emerging artemisinin resistance in Myanmar. Malar J. 2018;17(1):258. Epub 20180711. doi: 10.1186/s12936-018-2404-4. PubMed PMID: 29996844; PubMed Central PMCID: PMCPMC6042398.

41. Agyemang C, Boatemaa S, Agyemang Frempong G, de-Graft Aikins A. Obesity in Sub-Saharan Africa. In: (Ed.) RA, editor. Metabolic Syndrome A comprehensive textbook: Springer; 2016. p. 41–53.

42. Group NCDRFC-AW. Trends in obesity and diabetes across Africa from 1980 to 2014: an analysis of pooled population-based studies. Int J Epidemiol. 2017;46(5):1421–32. doi: 10.1093/ije/dyx078. PubMed PMID: 28582528; PubMed Central PMCID: PMCPMC5837192.

43. Ng M, Fleming T, Robinson M, Thomson B, Graetz N, Margono C, et al. Global, regional, and national prevalence of overweight and obesity in children and adults during 1980-2013: a systematic analysis for the Global Burden of Disease Study 2013. Lancet. 2014;384(9945):766–81. Epub 20140529. doi: 10.1016/S0140-6736(14)60460-8. PubMed PMID: 24880830; PubMed Central PMCID: PMCPMC4624264.

44. Griffin JT, Ferguson NM, Ghani AC. Estimates of the changing age-burden of Plasmodium falciparum malaria disease in sub-Saharan Africa. Nat Commun. 2014;5:3136. Epub 2014/02/13. doi: 10.1038/ncomms4136. PubMed PMID: 24518518; PubMed Central PMCID: PMCPMC3923296.

45. Obebe OO, Falohun OO. Epidemiology of malaria among HIV/AIDS patients in sub-Saharan Africa: A systematic review and meta-analysis of observational studies. Acta Trop. 2021;215:105798. Epub 2020/12/20. doi: 10.1016/j.actatropica.2020.105798. PubMed PMID: 33340524.

46. Walimbwa SI, Lamorde M, Waitt C, Kaboggoza J, Else L, Byakika-Kibwika P, et al. Drug Interactions between Dolutegravir and Artemether-Lumefantrine or Artesunate-Amodiaquine. Antimicrob Agents Chemother. 2019;63(2). Epub 2018/11/14. doi: 10.1128/AAC.01310-18. PubMed PMID: 30420479; PubMed Central PMCID: PMCPMC6355558.

47. Usman SO, Oreagba IA, Akinyede AA, Agbaje EO, Akinleye MO, Onwujuobi AG, et al. Effect of nevirapine, efavirenz and lopinavir/ritonavir on the therapeutic concentration and toxicity of lumefantrine in people living with HIV at Lagos University Teaching Hospital, Nigeria. J Pharmacol Sci. 2020;144(3):95–101. Epub 2020/09/15. doi: 10.1016/j.jphs.2020.07.013. PubMed PMID: 32921396.

48. Francis J, Barnes KI, Workman L, Kredo T, Vestergaard LS, Hoglund RM, et al. An Individual Participant Data Population Pharmacokinetic Meta-analysis of Drug-Drug Interactions between Lumefantrine and Commonly Used Antiretroviral Treatment. Antimicrob Agents Chemother. 2020;64(5). Epub 2020/02/20. doi: 10.1128/AAC.02394-19. PubMed PMID: 32071050; PubMed Central PMCID: PMCPMC7179577.

49. Muhindo MK, Kakuru A, Jagannathan P, Talisuna A, Osilo E, Orukan F, et al. Early parasite clearance following artemisinin-based combination therapy among Ugandan children with uncomplicated Plasmodium falciparum malaria. Malar J. 2014;13:32. Epub 20140128. doi: 10.1186/1475-2875-13-32. PubMed PMID: 24468007; PubMed Central PMCID: PMCPMC3909240.

50. Roberds A, Ferraro E, Luckhart S, Stewart VA. HIV-1 Impact on Malaria Transmission: A Complex and Relevant Global Health Concern. Front Cell Infect Microbiol. 2021;11:656938. Epub 20210412. doi: 10.3389/fcimb.2021.656938. PubMed PMID: 33912477; PubMed Central PMCID: PMCPMC8071860.

51. Saito M, Mansoor R, Kennon K, Anvikar AR, Ashley EA, Chandramohan D, et al. Efficacy and tolerability of artemisinin-based and quinine-based treatments for uncomplicated falciparum malaria in pregnancy: a systematic review and individual patient data meta-analysis. Lancet Infect Dis. 2020;20(8):943–52. Epub 20200429. doi: 10.1016/S1473-3099(20)30064-5. PubMed PMID: 32530424; PubMed Central PMCID: PMCPMC7391007.

52. Kloprogge F, Workman L, Borrmann S, Tekete M, Lefevre G, Hamed K, et al. Artemetherlumefantrine dosing for malaria treatment in young children and pregnant women: A pharmacokinetic-pharmacodynamic meta-analysis. PLoS Med. 2018;15(6):e1002579. Epub 2018/06/13. doi: 10.1371/journal.pmed.1002579. PubMed PMID: 29894518; PubMed Central PMCID: PMCPMC5997317 following competing interests: KIB and NJW are members of the WHO Technical Expert Group (TEG) on Malaria Chemotherapy. KIB is also a member of the WHO TEG on Drug Resistance and Containment. KIB, NJW, JT and SP are members of the WHO Malaria Chemotherapy sub-group on dosage recommendations. GL, KH, FE and RB are employees of Novartis, the manufacturer of the drug that is the subject of this publication. EAA and NJW are members of the Editorial Board of PLOS Medicine. None of the authors declare any other conflict of interest.

53. Onyamboko MA, Hoglund RM, Lee SJ, Kabedi C, Kayembe D, Badjanga BB, et al. A Randomized Controlled Trial of Three- versus Five-Day Artemether-Lumefantrine Regimens for Treatment of Uncomplicated Plasmodium falciparum Malaria in Pregnancy in Africa. Antimicrob Agents Chemother. 2020;64(3). Epub 2019/12/11. doi: 10.1128/AAC.01140-19. PubMed PMID: 31818818; PubMed Central PMCID: PMCPMC7038309.

54. Saito M, Gilder ME, Nosten F, McGready R, Guerin PJ. Systematic literature review and metaanalysis of the efficacy of artemisinin-based and quinine-based treatments for uncomplicated falciparum malaria in pregnancy: methodological challenges. Malar J. 2017;16(1):488. Epub 20171213. doi: 10.1186/s12936-017-2135-y. PubMed PMID: 29237461; PubMed Central PMCID: PMCPMC5729448.

